# Analysis of Influencing Factors of Conversion from Robot-Assisted Partial Nephrectomy to Radical Nephrectomy

**DOI:** 10.64898/2025.11.27.25341176

**Authors:** Baoling Zhang, Liwei Jing, Andi Wang, Jun Liu, Bin Han, Yanli Chai, Xin Gu

## Abstract

**Objective:** To analyze the influencing factors of conversion from robot-assisted partial nephrectomy (RAPN) to radical nephrectomy (RN).

**Methods:** The clinical data and surgical records of 258 patients who underwent RAPN at Cangzhou Hospital of Integrated Traditional Chinese and Western Medicine in Hebei Province from January 2021 to November 2025 were retrospectively collected. The patients were divided into the conversion group and the non-conversion group based on whether the operation was converted to RN intraoperatively. The relevant clinical data of the two groups were compared, and the reasons and influencing factors for conversion to RN were analyzed.

**Results:** Of the 258 patients, 8 (3.1%) were converted to RN. The reasons for conversion from RAPN to RN include: complicated tumor anatomy and technical difficulty; suspicion of locally advanced renal tumor intraoperatively; severe intraoperative bleeding. Univariate analysis showed that the R.E.N.A.L score (P<0.001), renal hilar tumor (P<0.001) and T stage (P=0.002) were the influencing factors for conversion to RN.

**Conclusion:** The main reasons for conversion from RAPN to RN are complex tumor anatomy, suspicion of locally advanced renal tumors intraoperatively, and severe intraoperative bleeding. The R.E.N.A.L score, renal hilar tumor location, and T stage are the influencing factors for conversion from RAPN to RN.

**Funding information:** Research Fund Project of the Hebei Provincial Health Commission (20240915)

## 1. Introduction

With the widespread application of robot-assisted technology in urology, robot-assisted partial nephrectomy (RAPN) has become one of the important surgical procedures for treating localized renal tumors due to its advantages of being minimally invasive, fast postoperative recovery, and a high nephron retention rate ^[1, 2]^. However, during the surgical process, it may be necessary to convert to radical nephrectomy (RN) for various reasons, which not only increases the risk of intraoperative trauma and postoperative complications for patients, but may also affect their prognosis ^[3]^. Therefore, clarifying the influencing factors for intraoperative conversion to RN is of great clinical significance for improving the success rate of surgery and the prognosis of patients. At present, there are relatively few studies on the influencing factors for intraoperative conversion from RAPN to RN. This study aims to explore the influencing factors for intraoperative conversion to RN by retrospectively analyzing the clinical data of patients who underwent RAPN in our institution, so as to provide a reference for clinical practice ^[4, 5]^.

## 2. Data and Methods

### 2.1 Study Population

This study’s retrospective design was approved by the institutional review board of Cangzhou Integrated Traditional Chinese and Western Medicine Hospital in Hebei Province. Data were accessed for research purposes on 1 November 2025.We retrospectively analyzed the clinical data of 258 patients who underwent RAPN at our institution from January 2021 to November 2025. Inclusion criteria were as follows: Patients diagnosed with renal tumors via preoperative imaging examinations (e.g., CT, MRI) and meeting the indications for RAPN; postoperative pathology confirmed the tumor as malignant or benign. Exclusion criteria were as follows: Patients who had been identified as requiring RN before surgery; patients with severe dysfunction of major organs (e.g., heart, lungs, liver, kidneys). Among these patients, 8 were converted to RN intraoperatively (conversion group), and 250 were not converted (non-conversion group). The patient records were anonymized and de-identified prior to analysis.

### 2.2 Variables

We collected clinical data of the two groups, including age, gender, body mass index (BMI), presenting symptoms (hematuria, low back pain, abdominal pain, incidental finding on medical examination, and others), personal history (tobacco use history and alcohol use history), comorbidities (hypertension, diabetes, coronary heart disease, cerebral infarction), history of abdominal surgery, preoperative creatinine level, preoperative eGFR, R.E.N.A.L score, tumor location, T stage, intraoperative blood loss, and tumor histologic type.

### 2.3 Statistical Analysis

Statistical analysis was performed using SPSS 25.0. Continuous data were expressed as median (M) and interquartile range (IQR), while categorical data were presented as counts and percentages. Continuous data were analyzed using the independent samples t-test; the Wilcoxon rank-sum test was used if the continuous data did not meet the t-test assumptions. Categorical data were analyzed using the chi-square test or Fisher’s exact test. P < 0.05 was considered statistically significant.

## 3. Results

### 3.1 Baseline Characteristics of the Study Subjects

This study included a total of 258 patients who underwent RAPN, with a median age of 55.6 years (range, 28–78 years), of whom 163 were male (63.2%) and 95 were female (36.8%). Eight patients (3.1%) were converted to RN, while the remaining 250 successfully underwent RAPN. There was no statistically significant difference (P > 0.05) between the two groups in terms of age, gender, BMI, presenting symptoms (hematuria, low back pain, abdominal pain, incidental finding on medical examination, and others), personal history (tobacco use history and alcohol use history), comorbidities (hypertension, diabetes, coronary heart disease, cerebral infarction), and other baseline data, which were comparable (Table 1).

### 3.2 Intraoperative conversion to RN

Among the 258 patients, 8 (3.1%) were converted to RN intraoperatively. The main reasons for conversion are as follows: Complex tumor anatomy and technical difficulty: 5 cases (62.5%), which manifested as tumors located near the renal hilum, closely associated with renal blood vessels, involving renal parenchyma in multiple planes, or having abnormal intrarenal vascular branches. These factors made it difficult to accurately dissect tumor tissue during RAPN. Conversion was performed to avoid serious postoperative complications such as urinary fistula and renal parenchymal ischemia. Suspected intraoperative locally advanced renal tumor: 2 cases (25.0%). Intraoperative exploration revealed tumor invasion of perirenal adipose tissue or suspected formation of a renal vein tumor thrombus. It was considered that the tumor stage might be T3 or higher, and thus the procedure was converted to achieve curative resection. Severe intraoperative bleeding: 1 case (12.5%). Severe bleeding occurred after incision of the renal parenchyma during surgery. Despite various hemostatic measures (e.g., suturing, application of hemostatic materials), bleeding could not be effectively controlled. RN was performed to ensure the patient’s safety.

### 3.3 Univariate analysis results

Univariate analysis was conducted on factors that might affect intraoperative conversion to RN, and the results showed the following: 1. R.E.N.A.L score: The R.E.N.A.L score serves as an important indicator for evaluating the anatomical complexity of renal tumors. Scores were divided into three groups: low-risk (4–6 points), moderate-risk (7–9 points), and high-risk (10–12 points). Patients in the conversion group had significantly higher R.E.N.A.L scores than those in the non-conversion group (P < 0.001). The conversion rate in the low-risk group was 1.7% (3/178), 5.3% (4/76) in the moderate-risk group, and 25.0% (1/4) in the high-risk group. The difference between groups was statistically significant (χ² = 21.345, P < 0.001). 2. Renal hilar location: Based on the relationship between the tumor and the renal hilum, tumors were classified as anterior renal hilar location, posterior renal hilar location, or complex location (involving both sides of the renal hilum or deep within the renal hilum). The proportion of tumors with complex renal hilar locations in the conversion group was significantly higher than that in the non-conversion group (P < 0.001). The conversion rate was 1.9% (2/108) for anterior renal hilar location, 2.3% (3/129) for posterior renal hilar location, and 14.3% (3/21) for complex location. The difference between groups was statistically significant (χ² = 18.762, P < 0.001). 3. T stage: According to the TNM staging system, the higher the T stage, the higher the conversion rate. The conversion rate was 1.4% (2/140) for T1a, 3.7% (4/108) for T1b, and 20.0% (2/10) for T2. The difference between groups was statistically significant (P = 0.002, Fisher’s exact test). 4. Comparison of tumor pathological types (P = 0.725) and intraoperative blood loss (P = 0.110) between the conversion and non-conversion groups showed no statistically significant difference.

## 4. Discussion

The conversion rate from RAPN to RN in this study was 3.1% (8/258), which is consistent with the previously reported conversion rate of 2%–5% ^[4, 6, 7]^. Although partial nephrectomy (PN) is the standard treatment for early-stage renal cell carcinoma and can preserve renal function to the greatest extent, the decision to convert to RN based on intraoperative real-time assessment of tumor anatomical complexity, invasion extent, and bleeding risk is an important measure to ensure curative tumor resection and patient safety ^[8, 9]^. In this study, the primary reason for conversion was complex tumor anatomy (62.5%), indicating that precise assessment of tumor location and renal vascular anatomy remains a core determinant in the surgical strategy for RAPN.

The R.E.N.A.L score and renal hilar location are key indexes reflecting tumor anatomical characteristics ^[10, 11]^. The R.E.N.A.L score quantifies surgical difficulty through parameters such as tumor size, location, and depth. This study shows that the score in the conversion group was significantly higher than that in the non-conversion group (P < 0.001), with a conversion rate of 25% in patients with high-risk scores (10–12). This is consistent with previous studies concluding that patients with high-risk scores have an increased risk of conversion from PN to RN ^[12]^. The conversion rate in patients with complex hilar locations (involving both anterior and posterior hilar regions or located deeply within the hilum) was 14.3%, which was significantly higher than that in patients with simple anterior or posterior tumors (1.9%–2.3%). This is closely related to the dense blood vessels in the hilar area and the difficulty in safe dissection when tumors invade renal pedicle branches ^[4]^. Difficulty in dissection due to anatomical complexity may prolong the duration of renal warm ischemia or increase the risk of postoperative complications. At this time, converting to RN is a reasonable choice to avoid serious complications such as postoperative renal ischemia and urinary fistula ^[13, 14]^.

T stage is an important index reflecting tumor invasion. In this study, the conversion rate of patients with T2 stage (20%) was significantly higher than that with T1 stage (1.4%–3.7%), suggesting that intraoperative judgment of local tumor progression is an important driving factor for conversion. Two cases were converted due to “suspected locally advanced renal tumor”, with intraoperative findings of perirenal fat invasion or suspected venous tumor thrombus, which is consistent with the 2022 EAU guidelines recommending that “relative contraindications for PN include clinical T3/T4 tumors” ^[15]^. It is worth noting that some T1 tumors are located deep in or close to the renal hilum; even with early clinical staging, these tumors may still be converted due to anatomical factors, suggesting that preoperative staging should be comprehensively evaluated in combination with anatomical scores ^[6]^. Although intraoperative frozen section pathology can assist in determining tumor nature, the evaluation of local invasion relies on the operator’s visual inspection, which has certain limitations. In the future, fluorescence imaging technology or molecular marker detection could be explored to improve evaluation accuracy ^[16, 17]^.

In this study, only one case (12.5%) was converted due to severe bleeding, which was lower than the bleeding-related conversion rate reported in the literature. This may be related to the precision of robotic surgical systems and advances in hemostatic techniques ^[4, 8]^. However, it is necessary to be alert that if main renal artery injury occurs after renal parenchymal incision, and simple suturing or hemostatic materials fail to control bleeding, timely conversion to RN is key to avoiding hemorrhagic shock ^[5]^. This suggests that Computed Tomography Angiography should be used preoperatively to accurately evaluate renal vascular anomalies, and priority should be given to managing renal artery branches intraoperatively to reduce the risk of main artery injury ^[18]^.

This study confirms that the R.E.N.A.L score, renal hilar location, and T stage are independent influencing factors for conversion. It is recommended that tumor anatomical features be accurately evaluated via 3D renal vascular reconstruction technology preoperatively. For patients with high-risk scores (≥10 points), complex renal hilar locations, and T2 stage tumors, the possibility of conversion should be fully informed to patients, and alternative surgical strategies should be formulated. Intraoperatively, the principle of “tumor cure takes priority over renal function preservation” should be followed, and timely decisions should be made for cases that are difficult to dissect or suspected of invasion ^[19]^.

The limitations of this study include: The sample size is small (only 8 conversion cases), and multivariate logistic regression analysis was not performed, which may overlook other potential influencing factors (e.g., surgeon experience and operative time); Intraoperative judgment of “locally advanced tumor” depended on the surgeon’s subjective evaluation and lacked confirmation with postoperative pathology; The long-term impact of conversion on postoperative renal function and tumor prognosis was not evaluated. In the future, it will be necessary to conduct multi-center, large-sample studies, and by combining postoperative pathological staging and survival data, to further clarify the optimal timing and clinical value of conversion decisions.

## 5. Conclusion

In summary, the main reasons for conversion from RAPN to RN are complex tumor anatomy, intraoperative suspicion of locally advanced renal tumors, and severe intraoperative bleeding. The R.E.N.A.L score, renal hilar location, and T stage are independent influencing factors for conversion from RAPN to RN. Clinicians should emphasize preoperative evaluation of tumor size and location, closely monitor intraoperative blood loss, and formulate a sound plan for conversion from RAPN to RN. Meanwhile, it is necessary to enhance surgeon training and refine surgical techniques to reduce the conversion rate and optimize patient prognosis.

## Data Availability

All relevant data are within the manuscript and its Supporting Information files.

## Conflict of Interest

All authors declare no conflict of interest.

## Author Contributions

BZ and XG were involved in conception and design, revising the article for intellectual content, and gave final approval of the completed version. LJ, AW, JL, and BH were involved in data extraction and drafting the article. All authors read and approved the final manuscript.

## Ethical Approval

All procedures performed in studies involving human participants were in accordance with the ethical standards of the institutional and/or national research committees and with the 1964 Helsinki Declaration and its later amendments or comparable ethical standards.

## Informed Consent

Informed consent was obtained from all individual participants included in the study.

## Consent for Publication

All authors agree to the publication of this article.

## Data Availability Statement

All data generated or analyzed during this study are included in this published manuscript.

## Notes

### Competing Interest Statement

The authors have declared no competing interest.

### Funding Statement

Yes

### Author Declarations

This study's retrospective design was approved by the institutional review board of Cangzhou Integrated Traditional Chinese and Western Medicine Hospital in Hebei Province.

